# Brain lesions causing parkinsonism versus seizures map to opposite brain networks

**DOI:** 10.1101/2024.05.02.24306764

**Authors:** Frederic L.W.V.J. Schaper, Mae Morton-Dutton, William Drew, Sanaz Khosravani, Juho Joutsa, Michael D. Fox

## Abstract

Recent epidemiological studies propose an association between parkinsonism and seizures, but the direction of this association is unclear. Focal brain lesions causing new-onset parkinsonism versus seizures may provide a unique perspective on the causal relationship between the two symptoms and involved brain networks. We studied lesions causing parkinsonism versus lesions causing seizures and utilized human connectome data to identify their connected brain networks. Brain networks for parkinsonism and seizures were compared using spatial correlations on a group and individual lesion level. Lesions not associated with either symptom were used as controls. Lesion locations from 29 patients with parkinsonism were connected to a brain network with the opposite spatial topography (spatial *r*=-0.85) compared to 347 patients with lesions causing seizures. A similar inverse relationship was found when comparing the connections that were most specific for lesions causing parkinsonism versus seizures on a group level (spatial *r*=- 0.51) and on an individual lesion level (average spatial *r*=-0.042; p<0.001). The substantia nigra was found to be most positively correlated to the parkinsonism network but most negatively correlated to the seizure network (spatial *r*>0.8). Brain lesions causing parkinsonism versus seizures map to opposite brain networks, providing neuroanatomical insight into conflicting epidemiological evidence.

## Introduction

In 1928, Yakovlev published an influential case series of individuals with epilepsy who developed parkinsonism.^1^ Yakovlev proposed that the neuroanatomical damage associated with life-long seizures may increase the risk of parkinsonism, suggesting shared pathophysiology. In contrast, Yakovlev also noted that in these cases, seizures decreased or even vanished after the onset of parkinsonism, proposing that the neuroanatomical damage associated with parkinsonism may protect against seizures.

Since Yakovlev’s clinical observations, recent epidemiological studies have observed a positive relationship between seizures and parkinsonism; having one diagnosis increases the chances of having the other.^2,3^ However, there is also evidence for an inverse relationship, as seizures can improve parkinsonism,^4,5^ and the onset of parkinsonism can improve seizures.^1,4^ Understanding the causal relationship between these two common brain diseases is potentially important for understanding pathophysiology, prognosis, and treatment. However, sorting out this relationship with epidemiology alone is difficult and may become more difficult over time with the increasing availability of treatment. For example, antiseizure drugs can induce parkinsonism symptoms and may increase the risk of Parkinson’s disease.^6^

Here, we first collate existing epidemiological data to assess whether the findings favor a positive or inverse relationship between parkinsonism and seizures. Next, we investigate this relationship from a unique perspective, examining cases where focal brain lesions cause new-onset parkinsonism versus cases where lesions cause new-onset seizures. If lesions causing these symptoms map to common neuroanatomy, that would suggest shared pathophysiology and support prior observations of a positive relationship. Conversely, if lesions causing these symptoms map to different neuroanatomy, this could support prior observations of an inverse relationship.^7^

## Materials and Methods

This study was carried out in accordance with the Declaration of Helsinki, approved by the institutional review board of the Brigham and Women’s Hospital, Boston, Massachusetts, and exempted from obtaining informed consent based on the secondary use of research data.

### Literature review

We performed a literature search using the keywords (“Parkinsonism”, “Parkinson’s disease”) and (“epilepsy”, “seizures”) to identify studies reporting on the relationship between parkinsonism and seizures. Published studies, study types, and main results are summarized in Table 1.

**Table 1.**
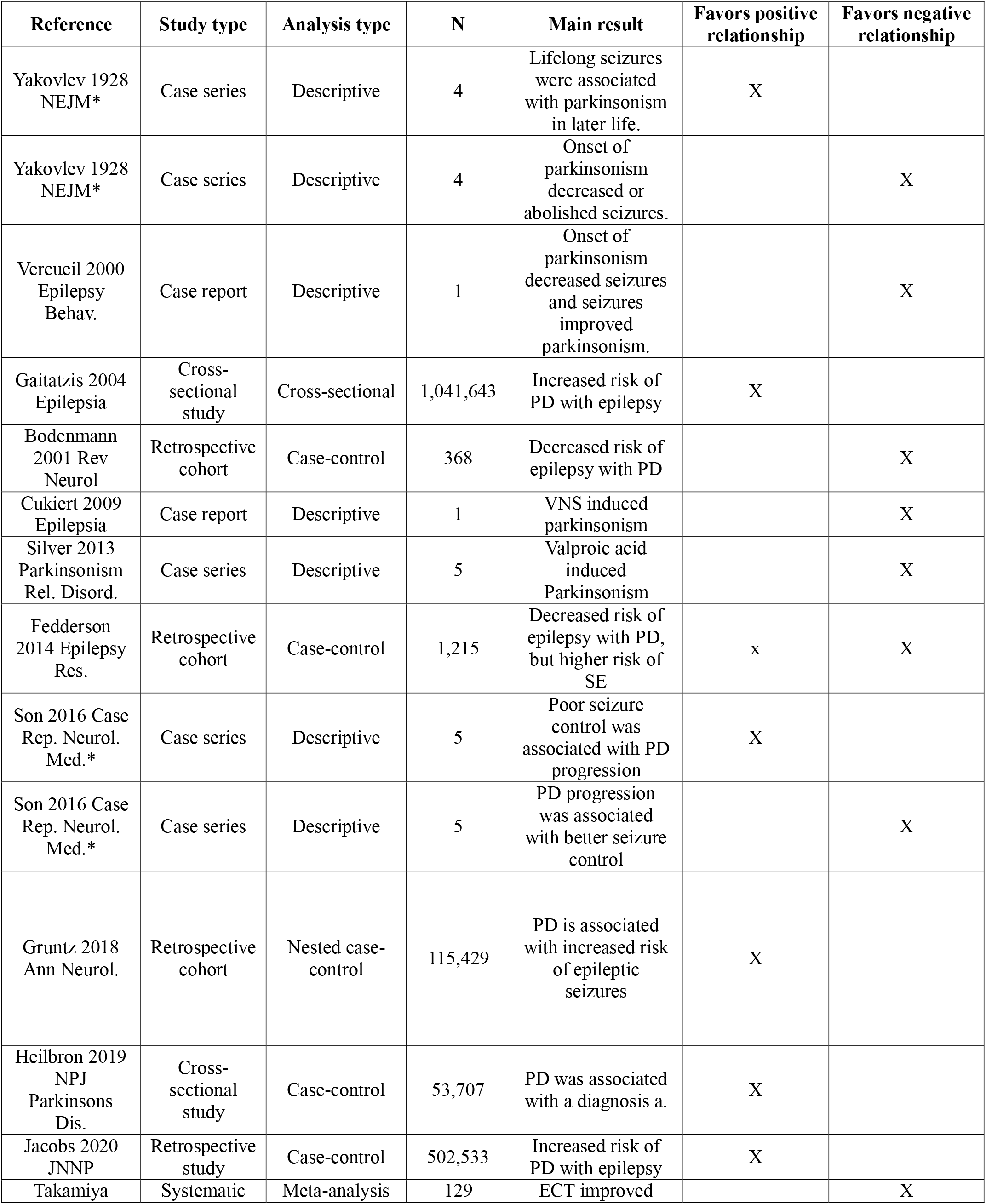
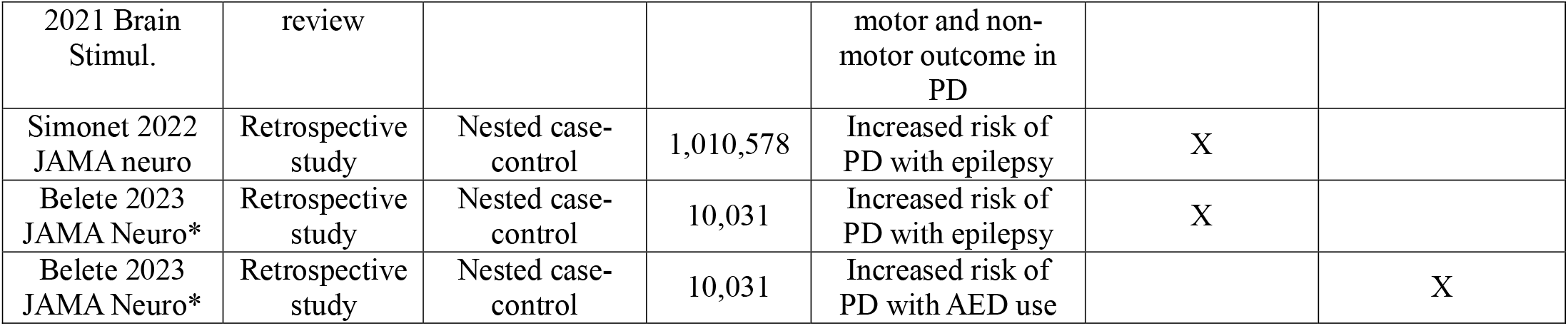
Literature summary on the relationship between parkinsonism and seizures. *Publications reporting results that favor both a positive and negative relationship.

### Lesions

Lesions were obtained from two previously published studies identifying brain networks for parkinsonism^8^ or seizures^9^. For the parkinsonism network, 29 lesions associated with parkinsonism and 135 lesions not associated with parkinsonism were studied.^8^ For the seizure network, 347 lesions associated with epilepsy and 1126 lesions not associated with epilepsy were studied.^9^ The human connectome was used to identify the brain network connected to lesions associated with parkinsonism or seizures, a validated technique termed ‘lesion network mapping.’^7^

### Parkinsonism versus seizure network

Brain networks for parkinsonism^8,10^ and seizures^9^ were published previously and shown in Fig. 1A and B. To avoid bias, we first compared the spatial similarity between the published parkinsonism (**Fig. 1A**) and seizure (**Fig. 1B**) networks using a spatial correlation (Pearson’s *r*). These networks were highlighted as the primary network in our previous publications but were derived differently. The parkinsonism map represents the mean connectivity profile of 29 lesions causing parkinsonism (**Fig. 1A**, see Supplementary Fig. 5 from Siddiqi *et al*.^10^). The seizure map represents the mean connectivity profile to subcortical nodes more functionally connected to 347 lesions causing epilepsy versus 1126 lesions not causing epilepsy (**Fig. 1B**, see Fig. 4A from Schaper *et al*.^9^). Second, to account for connections that may not be specific to parkinsonism or epilepsy, we also compared lesions causing either symptom to the control lesions of the original papers not associated with parkinsonism (n=135) or seizures (n=1,126) using a voxel-wise two-sample t-test. This voxel-wise comparison results in whole-brain specificity maps derived in the same manner. We then compared these group-level specificity maps using a spatial correlation (**Fig. 1C and D**). Third, we compared the network of each individual parkinsonism and seizure lesion to each other (**Fig. 1E and F**). Specifically, we assessed 1) the whole-brain connectivity profile of each parkinsonism (n=29) and control (n=135) lesion to each seizure lesion (n=347), averaging across all seizure lesions; and 2) the whole-brain connectivity profile of each seizure (n=347) and control (n=1,126) lesion to each parkinsonism lesion (n=29), averaging across all parkinsonism lesions. The average spatial correlations between parkinsonism, seizure, and control lesions were then compared using a t-test. A two-tailed p-value < 0.05 was considered significant. Finally, as performed in prior studies,^11^ we used the human connectome to identify voxels whose whole-brain connectivity profile was most similar to the published parkinsonism or seizure network (“network hubs”, **Fig. 2A and B**).

**Figure 1.**
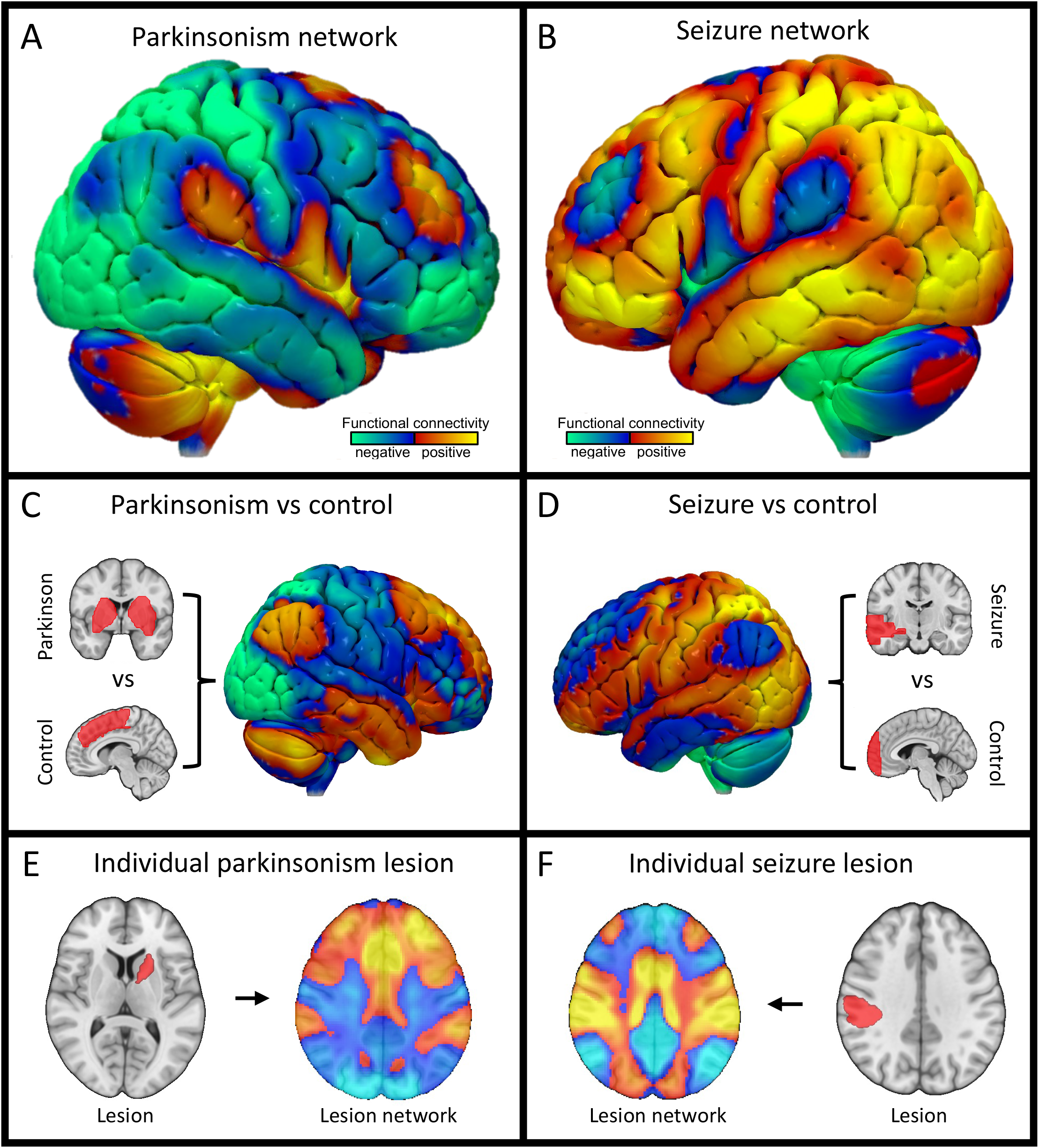
Brain lesions causing parkinsonism versus seizures map to opposite brain networks. We previously identified a parkinsonism network (**A**) and a seizure network (**B**) by combining lesions causing new-onset parkinsonism or seizure with an atlas of normative brain connectivity (the human connectome)^*7*^. First, we tested the spatial correlation between the published parkinsonism and seizure networks and found they were inversely related (spatial *r* = - 0.85). Second, we compared lesions causing parkinsonism to control lesions (**C**) and lesions causing seizures to control lesions (**D**) and found that these two group-level specificity maps also showed an inverse relationship (spatial *r* = -0.51). Third, we compared each individual parkinsonism and seizure lesion to each other and found a similar inverse relationship (average spatial *r* [95% CI] = -0.042 [-0.0484:-0.0360]; one-sample t-test p < 0.001).

**Figure 2.**
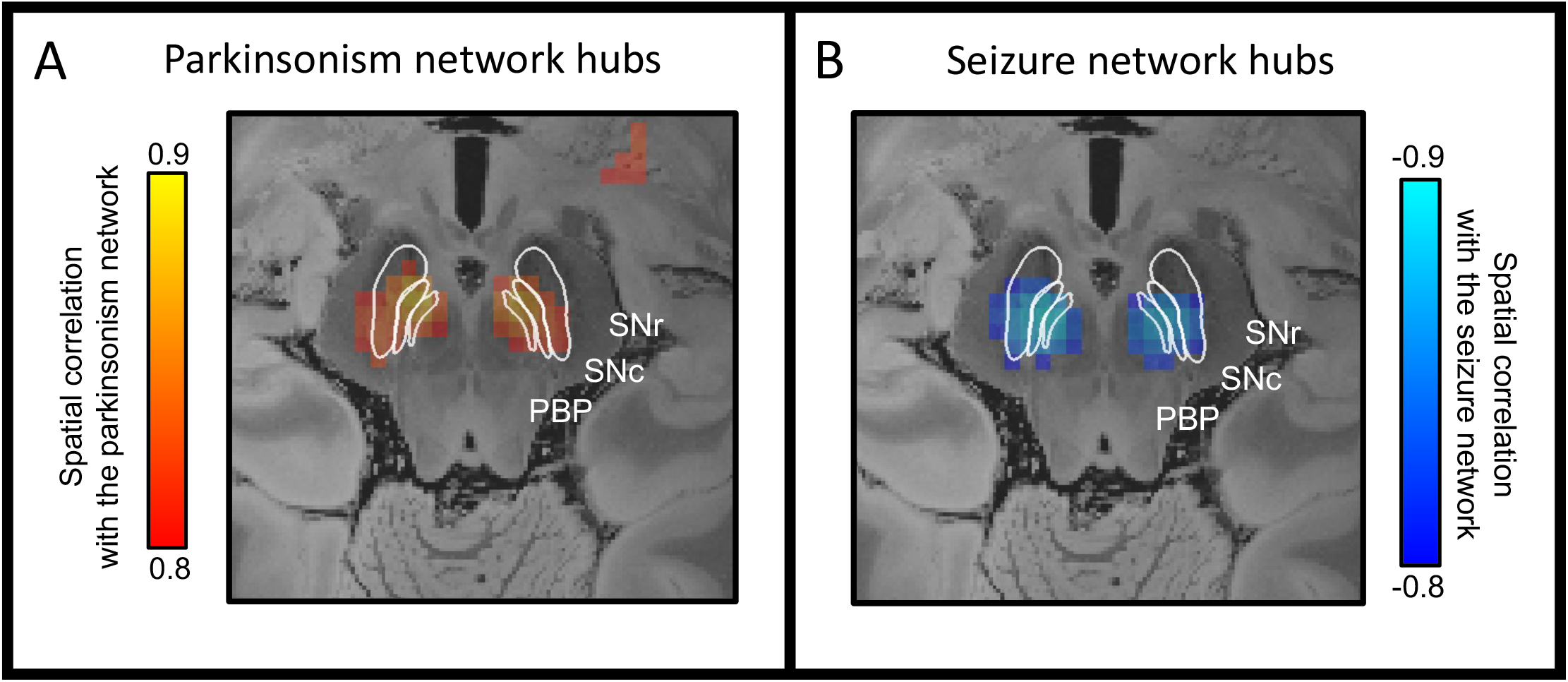
Parkinsonism and seizure network hubs. We identified the hubs in the parkinsonism and seizure network by computing the voxels whose whole-brain connectivity profile was most spatially similar to either the parkinsonism network (Fig. 1A) or the seizure network (Fig. 1B). Voxels in the substantia nigra were most positively correlated to the parkinsonism network (**E**) and most negatively correlated to the seizure network (**F**). *Abbreviations: SNr, substantia nigra pars reticulata; SNc, substantia nigra pars compacta; PBP, parabrachial pigmented nucleus*.

### Data availability statement

This paper used de-identified data from different teams of investigators at various institutions, across different countries. Each dataset is available upon reasonable request from each respective team of investigators. Data sharing will be subject to the policies and procedures of the institution as well as the laws of the country where each dataset was collected.

## Results

### Literature review

We identified 13 studies that reported on the relationship between parkinsonism and seizures. Of these studies, five supported a positive relationship between the two symptoms, five a negative, and three provided evidence for both (**Table 1**).

### Parkinsonism versus seizure network

Lesions causing parkinsonism are connected to a brain network (**Fig. 1A**) with the opposite spatial topography of lesions causing seizures (**Fig. 1B**, spatial *r* = -0.85). A similar inverse relationship (spatial *r* = -0.51) was found when computing the connections specific for lesions causing parkinsonism (**Fig. 1C**) or seizures (**Fig. 1D**). On an individual lesion level, the connectivity profile of lesions causing seizures showed a negative spatial correlation to the connectivity profile of lesions causing parkinsonism (average spatial *r* [95% CI] = -0.042 [- 0.0484 : -0.0360]; one-sample t-test p < 0.001, **Fig. 1E and F**). This negative spatial relationship was also specific compared to control lesions. Seizure lesions were more negatively correlated with parkinsonism lesions than control lesions not associated with parkinsonism (average spatial *r* [95% CI] = -0.004 [-0.035 : 0.028]; two-sample t-test, p = 0.018), and parkinsonism lesions were more negatively correlated with seizure lesions than control lesions not associated with seizures (average spatial *r* [95% CI] = 0.213 [0.183 : 0.242]; two-sample t-test, p < 0.001). On a brain network level, lesions causing parkinsonism are thus inversely related to lesions causing seizures.

Finally, we found that the network hubs (voxels with strongest positive or negative spatial correlation) in the published parkinsonism (**Fig. 1A**) and seizure network (**Fig. 1B**) were in the substantia nigra for both symptoms. However, the sign of the association was inverted. Voxels in the substantia nigra were most positively correlated to the parkinsonism network (**Fig. 2A**) and most negatively correlated to the seizure network (**Fig. 2B**).

## Discussion

In this study, we found that brain lesions causing parkinsonism and lesions causing seizures map to opposite brain networks, providing neuroanatomical insight into conflicting epidemiological evidence.^1,4^

Our results using brain lesions causing parkinsonism or seizures offer a unique perspective and suggest that parkinsonism and epilepsy are inversely related on a brain network level. This inverse relationship is consistent with multiple clinical and epidemiological findings of an inverse relationship, including reports of seizures improving parkinsonism^4,5^ and the onset or progression of parkinsonism improving seizures^1,4^ (see **Table 1**). Our results may also aid in interpreting prior epidemiological studies that observed a positive association^2,3^. Specifically, our results are consistent with the hypothesis that treatment of one disorder (not the presence of the disorder itself) may increase the risk of the other disorder, resulting in a positive association. Examples include antiseizure drug associated tremor and Parkinson’s disease.^3,6^ Mapping of lesions causing new-onset symptoms can allow for causal links between neuroanatomy and symptoms,^12^ akin to how Mendelian randomization allows for causal links between genes and symptoms.^13^

Regarding neuroanatomy, we found that the substantia nigra was a network hub in both the parkinsonism and seizure network, but the sign of the association was reversed. This means that the substantia nigra is the most likely location in the brain where a lesion would be expected to cause parkinsonism, consistent with the pathophysiology of Parkinson’s Disease. However, it is the least likely location for a lesion to cause seizures and could potentially improve or protect against seizures. This result is consistent with Yakovlev’s clinical observations in 1928,^1^ and results in experimental animals where direct lesioning, high-frequency stimulation, or optogenetic inhibition of the substantia nigra reduces seizures.^14^

Our results may have implications for side-effect monitoring during the development of new treatments. Treatments targeting the brain network of one disease could worsen the other. For example, epidemiological data suggest an increased risk of parkinsonism with common antiseizure drugs.^6^ Conversely, D1 selective medications are being explored as a more effective option for parkinsonism, but experimental data suggests these medications may confer a higher risk for seizures.^15^

